# A whole-brain modeling framework for tDCS montage optimization in drug-resistant epilepsy

**DOI:** 10.64898/2026.09.22.26363221

**Authors:** Borja Mercadal, Edmundo Lopez-Sola, Julia Makhalova, Francesca Pizzo, Ricardo Salvador, Fabrice Bartolomei, Giulio Ruffini

## Abstract

**Objective:** Transcranial direct current stimulation (tDCS) is a promising non-invasive therapy for drug-resistant epilepsy, but the montage optimization pipelines used to personalize it often rely on biophysical head models that capture only the induced electric field and ignore the network dynamics through which stimulation influences epileptogenic activity. We aimed to develop and evaluate a montage optimization framework that incorporates individualized network dynamics.

**Approach:** We introduce a framework based on a digital twin, or *neurotwin*, a personalized replica of a patient’s brain that couples a biophysical head model of the stimulation-induced field with a personalized whole-brain model (WBM) of seizure dynamics constrained by structural connectivity and intracranial recordings. We applied it to produce optimal montages in 12 patients with drug-resistant epilepsy, compared them with conventional biophysics-based optimization, and assessed the framework retrospectively against the clinical outcomes of six treated patients.

**Main results:** Because the neurotwin optimization suppresses simulated spread by construction, the informative result is the divergence between pipelines: the network-informed and field-based optimizations produced substantially different montages despite delivering comparable inhibitory fields at the epileptogenic zone, and montages with nearly identical field distributions could yield markedly different network outcomes within the model. A minimum-replacement-set analysis traced these differences to field changes at a few propagation-zone and off-target parcels rather than to the overall field. The parcels whose inhibition most reduced spread tended to occupy highly connected (hub) positions in the structural connectome, a robust but partial association indicating that network structure shapes, but does not fully determine, where stimulation is most effective. In the retrospective analysis, model-predicted spread reduction tracked clinical seizure-frequency change in the expected direction, though not significantly (*n* = 6).

**Significance:** We hypothesize that accounting for individualized network dynamics may improve seizure control, a hypothesis now being tested prospectively (NCT06334952).

**Highlights:**

- A neurotwin couples a biophysical head model with a personalized whole-brain model (WBM) for tDCS optimization.
- neurotwin and field-based optimization yield divergent montages despite comparable EZ fields.
- Field differences at a few target and off-target parcels, not overall field magnitude, drive the simulated network outcomes.
- Parcels controlling simulated seizure spread tend to be structural network hubs (high nodal strength and centrality), an association that persists after adjusting for local dynamics.
- WBM-predicted spread reduction was directionally consistent with clinical seizure-frequency change (non-significant, *n* = 6).

## 1 Introduction

Transcranial direct current stimulation (tDCS) is a non-invasive neuromodulation technique in which weak direct currents, typically 1–2 mA, are delivered through scalp electrodes to modulate cortical excitability in a polarity-dependent manner.^1, 2^ By inducing subthreshold membrane potential perturbations in neurons, tDCS can alter spontaneous firing rates^3, 4^ and promote lasting neuroplastic changes.^5^ Due to its potential to non-invasively modulate brain networks, tDCS (and other variants of transcranial stimulation) has been applied in clinical trials to treat diverse neurological and psychiatric disorders, including depression, chronic pain, stroke, and epilepsy.^6, 7^

tDCS offers an attractive therapy for patients with drug-resistant epilepsy, due to its potential to reduce cortical hyperexcitability and alter epileptogenic network dynamics.^8, 9^ Early pilot studies applying cathodal tDCS over seizure onset zones demonstrated reductions in seizure frequency and interictal epileptiform discharges.^10, 11^ More recent clinical studies have adopted a personalized multi-channel approach in which subject-specific electrode configurations are designed based on anatomical and electrophysiological data. Open-label and sham-controlled studies have been performed using such an approach, with some patients achieving clinically meaningful seizure reductions.^12–16^ Overall, however, the evidence base remains limited and the reported responses heterogeneous. Most previous studies are small, several are open-label or uncontrolled, and a substantial fraction of patients show little or no benefit, motivating efforts to better understand and predict the treatment response.

Typically, personalized multi-channel tDCS relies on a montage optimization process that aims to select the electrode positions and current intensities that maximize stimulation at target regions while minimizing off-target effects.^17–19^ State-of-the-art optimization pipelines employ subject-specific biophysical head models derived from structural MRI, combined with finite element modeling, to predict the intracranial electric field distribution and solve an optimization problem based on predefined target maps.^17, 20^ While these methods offer high spatial precision in field delivery, they do not directly account for the network dynamics through which stimulation likely influences epileptogenic activity.

Over the past two decades, computational neuroscience has made major advances in modeling the large-scale brain network dynamics, integrating anatomical, physiological, and functional data into unified simulation frameworks.^21–23^ Platforms such as *The Virtual Brain* have demonstrated how individualized structural connectomes, derived from diffusion MRI, can be coupled to biophysically grounded neural mass or mean-field models to reproduce empirically observed functional connectivity and network dynamics.^24, 25^ Such models are increasingly used to mechanistically represent neurological disorders by embedding disorder-specific pathophysiological mechanisms into their equations. Examples include the alteration of excitation–inhibition balance in epilepsy,^26, 27^ dopaminergic deficits in Parkinson’s disease,^28^ and amyloid- and tau-related synaptic dysfunction in Alzheimer’s disease.^29, 30^

In epilepsy, brain network models have been used to delineate the epileptogenic zone (EZ) and propagation zones (PZ) from multimodal imaging and electrophysiology,^31–33^ to predict surgical outcomes^34^ and virtual-resection effects,^35–38^ and to interpret intracranial and scalp EEG by linking signals to network mechanisms.^39–42^

These frameworks can also serve as *in silico* testbeds for therapeutic interventions. Because their parameters map onto interpretable physiological processes, these models can simulate the effects of tDCS.^43^ Prior work, however, has mostly used them to simulate predefined stimulation protocols, to demonstrate *in silico* that seizure propagation can in principle be controlled by perturbing network nodes,^44^ or to optimize surgical interventions such as virtual resections.^35, 38^ More recently, stimulation has been embedded in personalized brain models, but primarily to estimate the epileptogenic network for diagnosis^45^ rather than to design therapy.

Here we present a *neurotwin*-based pipeline to optimize multichannel tDCS in drug-resistant epilepsy. A *neurotwin* is a personalized digital replica of a patient’s brain that can simulate the network response to stimulation and guide montage design. The *neurotwin* integrates two complementary modeling layers. The first is a biophysical head model, built from individual anatomical MRI, that predicts the electric field induced in the cortex by transcranial stimulation. The second is a personalized whole-brain model (WBM), implemented with the framework introduced by Lopez-Sola et al.,^46^ which couples patient-specific structural connectivity from diffusion MRI with physiologically grounded neural mass models tuned to intracranial recordings. These two layers are coupled by mapping the field predicted by the head model onto membrane-potential perturbations of the neural masses, so that any candidate montage yields a simulated network response. This coupling is what makes the pair a single replica rather than two models run in sequence. While previous studies used brain network models to predict the response to a predefined montage or estimate the epileptogenic network, in our pipeline the WBM becomes the objective that drives the optimization itself, closing the loop between simulation and treatment design. To the best of our knowledge, this is the first study to use a brain network model embedded in a biophysical model to optimize tDCS montages.

In this paper, we evaluate our neurotwin-based pipeline in a cohort of 12 patients and compare the montages it produces with those of a conventional biophysics-based optimization. We then use the WBM to dissect why the two approaches diverge and to identify the cortical regions responsible. Finally, we assess the framework retrospectively against the clinical outcomes of six previously treated patients.

## 2 Methods

The neurotwin pipeline for optimizing tDCS montages integrates personalized WBMs and biophysical head models. Figure 1a shows a schematic of the full pipeline.

**Figure 1:**
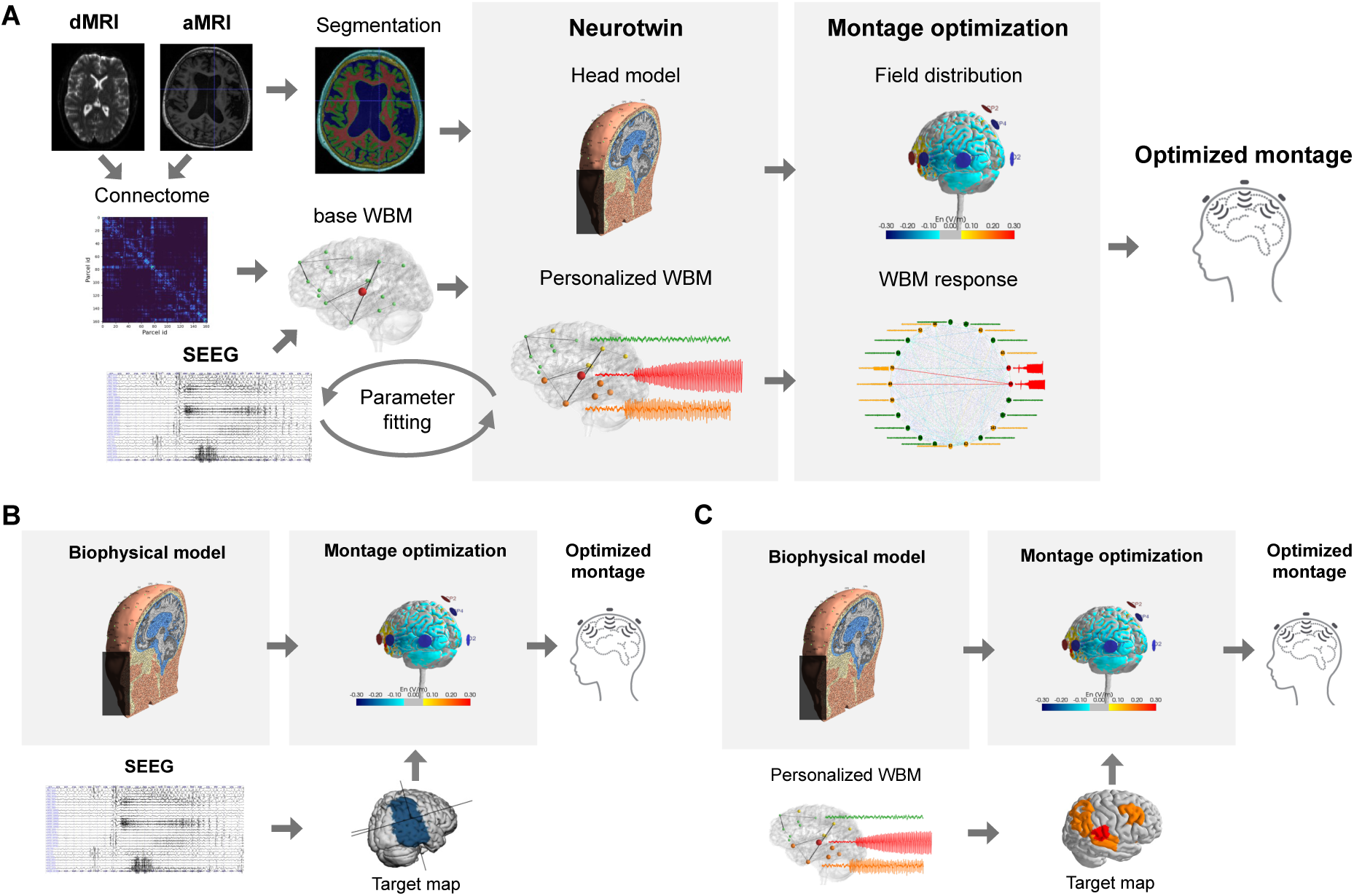
Optimization pipelines. a) Schematic of the neurotwin optimization pipeline presented in this article. Using imaging and SEEG data, a neurotwin is built, consisting of a personalized biophysical model and a personalized WBM. Then, montage optimization is done based on the field distribution and the WBM response. b) Optimization pipeline based on the biophysical model. SEEG data is used to define a target map and then optimization is done based on the field distribution. c) Optimization pipeline based on the biophysical model with the target map defined with the personalized WBM.

To analyze the optimization pipeline outputs, they were compared with those generated by physics-based optimization strategies. On the one hand, montages were generated following an established optimization approach^12–16^ (Figure 1b). A target map of the brain areas to inhibit was defined from the clinician’s assessment of the patient’s data, and an optimization algorithm found the montage that maximized inhibition of those regions while sparing others.

To assess the impact of the WBM in the neurotwin optimization pipeline, we used a hybrid approach in which physics-based optimization was applied using target and weight maps derived from WBM simulations (Figure 1c).

### 2.1 Data acquisition

In the present study, data from 12 patients, acquired as part of completed clinical studies targeting drug-resistant epilepsy in the context of the [GALVANI](https://cordis.europa.eu/project/id/855109/results) project (NCT04782869, NCT05250713)^13–16^ was used. Among participants in these trials, we selected patients with high-resolution anatomical MRI, diffusion MRI (dMRI), and stereo-electroencephalography (SEEG) recordings during spontaneous seizures. The clinical characteristics of the selected patients are summarized in Table 1.

**Table 1:**
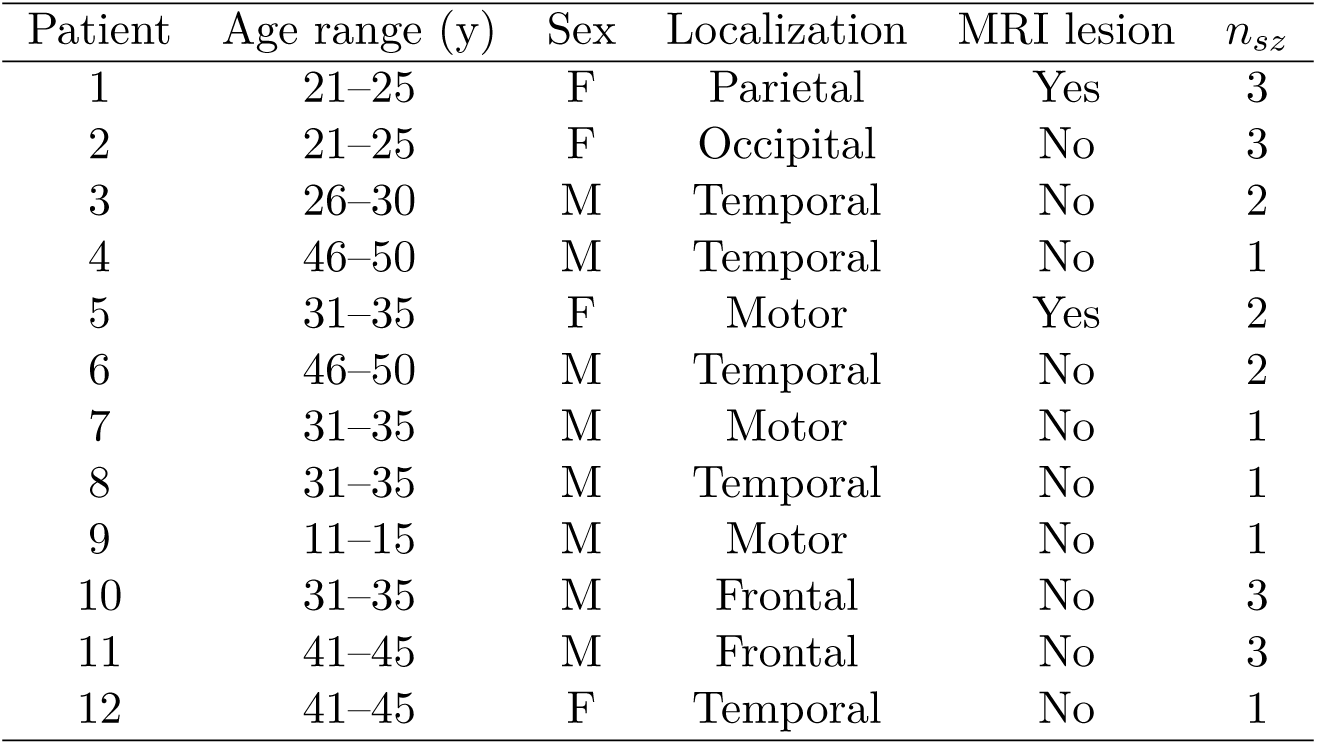
Clinical characteristics of the 12 patients. Localization indicates the seizure-onset (target) region. MRI lesion indicates whether a structural lesion was identified on anatomical MRI. *n_sz_* is the number of recorded seizures used to personalize the WBM. Ages are reported as non-overlapping 5-year ranges (years). n/a = not available.

SEEG implantation and clinical classifications of epileptogenic zones (EZ), propagation zones (PZ), and non-involved zones (NIZ) were performed by expert clinicians as part of the routine pre-surgical evaluation. All patient data acquisition procedures adhered to ethical standards and received approval from institutional review boards. Written informed consent was obtained from all participants in accordance with the Declaration of Helsinki. The secondary use of these data for the present modeling study was covered by the original study approvals and participant consent.

### 2.2 Data processing

Each patient’s T1-weighted MRI was parcellated into 162 cortical regions according to the Virtual Epileptic Patient (VEP) atlas^47^ using Freesurfer^48^ and custom Python scripts. Diffusion MRI images were processed to calculate the subject’s structural connectivity. Preprocessing steps included denoising, artifact correction (e.g., eddy currents), and estimating the fiber orientation density using constrained spherical deconvolution. Probabilistic tractography was then performed with MRtrix3^49^ generating 20 million streamlines constrained to start and terminate in gray matter parcels defined by the MRI parcellation. Finally, the connectivity matrix was obtained by quantifying the number of streamlines connecting each pair of parcels.

SEEG electrode locations were mapped to cortical parcels defined in the VEP atlas^47^ using the GARDEL software suite.^50^ Then, clinician interpretation of SEEG recordings was used to classify brain regions according to their role in seizure generation and spread. Specifically, each parcel in the VEP atlas was assigned to one of three functional categories: epileptogenic zone (EZ), propagation zone (PZ), or non-involved zone (NIZ).

### 2.3 Whole-brain Model Personalization

Subject-specific WBMs of seizure spread were built following the WBM personalization framework introduced by Lopez-Sola et al.^46^ A self-contained description of this procedure, covering the node models, the fitted and fixed parameters, the empirical and simulated functional connectivity, and the fitting and validity criteria, is provided in the Supplementary Methods on whole-brain model personalization. Briefly, a neural mass model (NMM) was placed in each region of the VEP parcellation atlas, with the specific model type depending on its classification.

For parcels labeled as EZ, a seizure-generating model developed by Lopez-Sola et al.^51^ was used, which incorporates chloride ion dynamics to simulate transitions from baseline to ictal states. For all other regions, including PZ and NIZ, a modified version of the Wendling-class model^39^ was used, in which seizure susceptibility is controlled by a region-specific excitatory synaptic gain parameter (*W*_exc_). These models were coupled following the individualized structural connectome, scaled by a global coupling parameter (*G*) that modulates the overall strength of long-range interactions.

Synthetic SEEG recordings were simulated following the laminar neural mass modeling framework^52^ and the biophysical SEEG model introduced in.^53^ These synthetic SEEG signals were used to compute functional connectivity matrices via amplitude envelope correlations, mirroring the approach used for the empirical SEEG data. The global coupling (*G*) and local excitability (*W*_exc_) for each SEEG-sampled parcel were then optimized to maximize the Pearson correlation coefficient between the simulated and empirical FC matrices. Per-patient model characteristics (the number of SEEG-sampled parcels, the EZ and PZ parcel counts, the achieved FC fit, and the number of *model propagation-zone* (mPZ) parcels, non-EZ parcels that seize in the model without stimulation) are summarized in Table 2.

**Table 2:** Per-patient WBM characteristics. SEEG parcels: number of VEP parcels sampled by SEEG contacts. EZ / PZ: number of parcels classified as epileptogenic zone and propagation zone, respectively. FC fit: Pearson correlation coefficient between the empirical and simulated functional-connectivity matrices. FC vs SC: Pearson correlation coefficient between the empirical functional connectivity and the structural connectome, a validity baseline that the FC fit must exceed.^46^ mPZ: number of non-EZ parcels exhibiting seizure activity in the model in the absence of stimulation (the model propagation zone).

| Patient | SEEG parcels | EZ | PZ | FC fit | FC vs SC | mPZ |
| --- | --- | --- | --- | --- | --- | --- |
| 1 | 23 | 2 | 3 | 0.72 | 0.27 | 9 |
| 2 | 22 | 2 | 0 | 0.58 | 0.13 | 4 |
| 3 | 40 | 6 | 3 | 0.58 | 0.07 | 18 |
| 4 | 28 | 2 | 6 | 0.89 | 0.27 | 44 |
| 5 | 18 | 6 | 2 | 0.64 | 0.05 | 5 |
| 6 | 33 | 5 | 5 | 0.84 | 0.23 | 148 |
| 7 | 12 | 4 | 4 | 0.82 | 0.43 | 5 |
| 8 | 44 | 6 | 17 | 0.66 | 0.10 | 135 |
| 9 | 32 | 2 | 16 | 0.38 | 0.14 | 14 |
| 10 | 25 | 4 | 7 | 0.37 | 0.21 | 155 |
| 11 | 25 | 2 | 5 | 0.83 | 0.33 | 153 |
| 12 | 37 | 4 | 9 | 0.76 | 0.19 | 137 |

We assessed model validity using the benchmark introduced with this personalization framework:^46^ a model was retained only if the correlation between its simulated and the empirical FC exceeded the correlation between the empirical FC and the structural connectome, confirming that the fitted model captures functional dynamics beyond what is already encoded in the anatomy. All 12 models met this criterion (Table 2), including the two with the lowest fits (patients #9 and #10, FC fit 0.38 and 0.37 against structural baselines of 0.14 and 0.21). The absolute FC fits are modest in several patients, which we attribute to the near-binary nature of the model FC: seizure detection assigns each parcel an all-or-none ictal state, so the simulated FC takes near-binary values, whereas the empirical FC is graded. This mismatch in dynamic range caps the achievable correlation even for a well-personalized model, so a moderate FC fit that still clears the structural baseline is expected rather than a sign of poor personalization. The number of mPZ parcels also varies widely across the cohort, from 4 to 155. This reflects two factors: within a patient many parcels can be strongly coupled to the EZ and are therefore recruited together once a seizure spreads, and parcels not sampled by SEEG do not enter the FC fit, so their simulated seizure activity is neither constrained nor penalized during personalization and can inflate the baseline spread.

### 2.4 Biophysical head models

For each patient, a personalized biophysical head model was produced from T1-weighted MRI images. First, SimNIBS 3.2.4^54^ *headreco* command was used to segment the MRI into 5 tissue types: scalp, skull (both spongy and compact bone were classified as one single tissue), cerebrospinal fluid (CSF, including the ventricles), gray-matter (GM) and white-matter (WM). All segmentations were inspected and corrected manually if deemed necessary. Then, SimNIBS was also used to build smooth triangulated surfaces from the segmentation masks that subsequently allowed creating a finite element volumetric mesh comprised of tetrahedra.

The 10–10 EEG system positions were identified on the scalp surface of the biophysical head model and the lead-field matrix in node space, 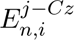 for the tES scalp electrodes was calculated. This is a matrix that contains the value of the electric field component normal to the cortical surface, *E_n,i_* for each coordinate in the triangulated cortical surface, *i*, for every possible bipolar montage of tES electrodes, *j* with one common cathode (Cz). This matrix can then be used to calculate the electric field generated by any montage using the electrode positions included in the lead field matrix (a detailed explanation can be found elsewhere^17^):

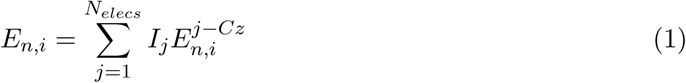

The electric field distribution for each pair of tES electrodes was computed with the finite element method in SimNIBS.^54^ The conductivity of the tissues was assumed isotropic and homogeneous. The values for each tissue were selected based on studies available in the literature: 0.33 S/m for the scalp, 0.008 S/m for the skull, 1.79 S/m for the CSF, 0.40 S/m for the GM and 0.15 S/m for the WM.^55–57^ Regarding the electrodes, only the gel needs to be represented geometrically, and this was done by creating a 3 mm thickness cylinder (with the same radius as the PiStim electrodes from Neuroelectrics) located at each of the previously identified scalp positions with a conductivity of 4 S/m.

The cortical surface parcellation from Freesurfer was mapped to the biophysical head model GM surface by finding the nearest neighbor between the surface meshes and correcting for disconnected components. Then, the leadfield matrix in parcel space, 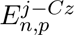, was calculated by averaging 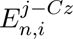 over all nodes *i* belonging to each parcel *p*. By doing so, the average electric field in a parcel induced by a montage can be calculated with Eq. 1 using 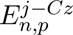 as a leadfield.

### 2.5 Biophysical model-based optimization

The Stimweaver algorithm^17^ was used to maximize the Normalized Error with respect to No Intervention, *NERNI*, calculated from the field distribution generated by a montage *E_n_* as:

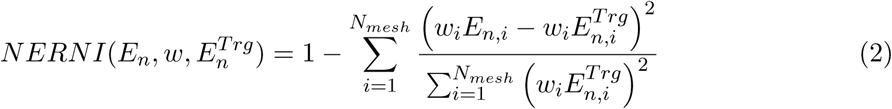

where *E_n,i_* is the normal component of the electric field in node *i* of the biophysical head model mesh on the gray matter surface and can be calculated with Eq. 1. *w_i_* and 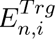 are the weight and the target field at node *i* respectively. Throughout this manuscript we will refer to the full vector of values of *w_i_*and 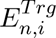 as the weight map and the target map.

To define the target and weight maps, we followed an approach intended to replicate that of previous clinical trials.^12–14, 16^ First, Neuroelectrics’ [online target editor](https://www.neuroelectrics.com/products/research/target-editor) was used to select the cortical areas to inhibit following the clinician’s assessment of the EZ and PZ regions (Supplementary Figure S1 shows an example for one patient). Then, using this information, a target field of 0.25 V/m and a weight of 20 was assigned to the mesh nodes in the regions to inhibit and a target field of 0 to the others. The weight in the nodes was calculated based on the focality ratio, *R*, defined as the ratio between the squared sum of weights in the target area and the squared sum of the weights outside of the target area.

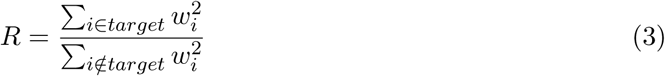

The weights in the nodes outside the target area were adjusted on a case-by-case basis to keep a constant ratio of 7.3 across all subjects. This value was adjusted heuristically to balance focality against field on target.

### 2.6 Neurotwin-based optimization

The optimization strategy is based on the *imprint hypothesis*: stimulation protocols are ranked by the desired acute change they induce in the target neural dynamics, under the assumption that repeated induction of this state biases activity-dependent plasticity toward a durable reorganization in the same functional direction. This hypothesis is supported by evidence that transcranial stimulation can engage NMDA- and BDNF-dependent plasticity and produce LTP-like and metaplastic effects lasting several hours, although the general correspondence between acute network modulation and long-term clinical benefit remains to be established.^5, 58, 59^

Within our modeling pipeline, we therefore assume that the acutely simulated neural response to stimulation, such as changes in seizure propagation or probability, is a valid proxy for the potential of a given montage to elicit the desired plastic effects through tDCS. We acknowledge, however, that the actual clinical efficacy of the intervention is influenced by additional factors not captured in the present framework, including stimulation session duration, inter-session interval, and the total number of sessions administered.

#### 2.6.1 Simulations of the electric field effects on the brain model

To simulate the effects of transcranial electrical stimulation (tES) in the WBM, the distribution of the electric field was first estimated and then mapped to membrane potential perturbations in the neural populations. For each brain parcel, the average normal component of the electric field, *E_n,p_*, induced by a given tES montage, was computed using the leadfield matrix in parcel space, 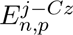. Subsequently, the membrane potential perturbations, *u_i,p_*, for population *i* in parcel *p*, were calculated as follows:

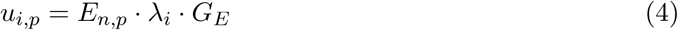

Here, the coefficients *λ_i_* were assigned based on the morphology and orientation of each neural population, with values derived from multi-compartmental modeling work.^60^ In this study, values of 1, 0.35, 0.25, and 0.1 were used for pyramidal cells, excitatory interneurons, SST interneurons, and PV interneurons, respectively. *G_E_* combines two contributions: a geometric factor that compensates the cancellation incurred when the signed normal field is averaged over the folded cortical surface of a parcel (necessary because the sensitivity coefficients*λ_i_* are defined for the local field at the neuron), and a mesoscopic amplification factor reflecting recurrent network coupling. As detailed in the next section, *G_E_* calibration absorbs both into a single per-subject value.

The computed perturbations, *u_i,p_*, were then used in the WBM by adding them to the membrane potential of the corresponding neural population in each parcel. Simulations were subsequently run as described in,^46^ and seizure spread was assessed using the internal variables in the model. Specifically, seizure activity in a parcel was detected if the firing rate of the excitatory interneuron population displayed more than one saturation peak. Throughout, the non-EZ parcels that exhibit such seizure activity at baseline (i.e., without stimulation) are the mPZ parcels, and the remaining non-EZ parcels are *model non-involved* (mNI) parcels. These model-defined categories reflect the seizure spread reproduced by the network model and need not coincide with the clinically defined PZ and NIZ. In practice, however, the two largely agree: across the cohort, 82% of clinically defined PZ parcels were also classified as mPZ.

#### 2.6.2 Electrical coupling gain adjustment

The electrical coupling gain, *G_E_*, accounts for the fact that the parcel-averaged membrane-potential perturbations induced by tES at the single-cell level are individually too small to drive a neural mass model, which represents the mean activity of a large population rather than single cells. Single-cell field sensitivity must therefore be scaled up to the population level, and this rescaling has a mechanistic basis: cancellation from signed-averaging *E_n_* over parcels and recurrent network coupling which amplifies weak per-cell perturbations into population-level effects.^61–64^ Consequently, it is common practice in neural mass models to introduce a scaling factor (the coupling gain) for such collective amplification. However, since this mesoscopic factor cannot be derived from microscopic considerations with realistic neurons, the scaling factor was instead adjusted so that the models respond to stimulation under physically realistic conditions.

Calibration of the coupling gain, *G_E_*, was performed on a per-subject basis by analyzing the response of the subject-specific model to single parcel targeted stimulation. For each mPZ parcel, a binary search algorithm was used to identify the minimum inhibitory electric field required to prevent seizure onset when stimulation was applied to that single parcel. Across all parcels, the maximum required field, *E_max_*, was selected as a reference value.

The coupling gain, *G_E_*, was then calculated so that this reference field corresponded to a value of 0.1 V/m, typical of the cortical fields generated by tDCS as predicted by models^65^ and verified experimentally:^66, 67^

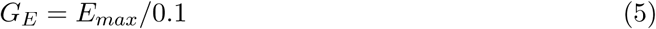

Anchoring the coupling gain to the hardest parcel to influence (the one requiring the largest field to suppress seizure activity) incorporates inter-individual and regional variability in tES responsiveness while keeping the model’s sensitivity consistent with experimentally observed field magnitudes. This 0.1 V/m reference is a *raw* cortical field, of the magnitude typically generated by tDCS. The quantity that actually drives the WBM, however, is the signed parcel-average normal field, which is much smaller because averaging the normal component over the folded cortical surface within a parcel produces substantial cancellation (across our cohort its absolute value had a median of 0.013 V/m and never exceeded 0.13 V/m). Requiring the hardest-to-suppress parcel to reach a raw field of 0.1 V/m is therefore a conservative calibration. Montages nonetheless suppress the relevant parcels because suppression is a network effect rather than a per-parcel field threshold.

#### 2.6.3 Optimization algorithm

Montage optimization was done by minimizing a loss function that has two main components, one derived from WBM simulations and another that is based on the biophysical head model. Formally, it is defined as:

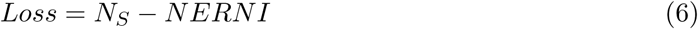

where *N_S_* is the number of non-EZ parcels with seizure in the NMM simulation when applying stimulation with the montage being evaluated. *NERNI* is the Normalized Error with respect to No Intervention calculated as in Eq. 2. Because *N_S_* is integer-valued and *NERNI* is bounded above by 1, montages are ranked primarily by the number of suppressed parcels, and the *NERNI* term acts only as a secondary criterion that breaks degeneracy. Since the montage space is continuous while *N_S_* takes only integer values, many distinct montages share the same *N_S_*, so the *NERNI* term is in practice always the operative criterion that selects among these equally-spreading montages the one with the most focal inhibitory field. The target maps and weight maps used in the NERNI calculation were defined as follows:

- For EZ and mPZ parcels, a target electric field of 0.25 V/m was set, while a target electric field of 0 V/m was set to the rest of the brain.
- For EZ parcels a weight of 10 was defined
- For mPZ parcels a weight of 2 was defined
- for the rest of parcels, the weight was adjusted using Eq. 3 with a focality ratio of 7.3

To find optimal montages, Eq. 6 was minimized using the differential evolution algorithm,^68^ specifically the implementation provided by the SciPy Python library using the best1bin strategy.^69^ The algorithm was run with a mutation constant sampled in the range (0.2, 0.6), a recombination constant of 0.4, a population size of 75 times the number of free parameters, and a maximum of 60 generations. The montage search space comprised the 64 electrode positions of the 10–10 EEG system, and candidate montages were constrained to be physically and clinically realizable: a maximum of 6 active electrodes, a total injected current of 2 mA, a maximum of 1.8 mA at any single electrode, and a minimum of 200 *µ*A at any active electrode. Because the optimizer searches only over montages satisfying these constraints, every returned solution is by construction deliverable within the same safety envelope.

### 2.7 Single-parcel inhibition analysis

To characterize how the personalized WBM responds to focal inhibition, we systematically stimulated one parcel at a time. For each parcel exhibiting seizure activity at baseline, an inhibitory perturbation was applied to that parcel alone and its magnitude increased until the local seizure was abolished, while all other parcels were left unstimulated. We then recorded which of the remaining parcels continued to seize. Repeating this for every seizing parcel yielded, for each subject, a *spread matrix* whose rows index the stimulated parcel and whose columns index the parcels in which seizure activity was observed. For each stimulated mPZ parcel we quantified the *spread reduction*: the fraction of baseline-seizing non-EZ parcels whose seizure was abolished when that parcel alone was inhibited.

To test whether the structural connectome shapes which parcels have the largest effect, we related the single-parcel spread reduction to standard graph-theoretical centrality metrics of each parcel. We computed three standard centrality measures on the weighted connectome: nodal strength (the sum of a parcel’s connection weights), eigenvector centrality, and betweenness centrality.^70^ As potential confounds for the structural association, we also recorded two descriptors of each parcel’s dynamical state in the personalized model: its fitted excitability parameter (*W*_exc_) and its baseline proximity to seizure onset, defined as the minimum inhibitory field required to abolish its seizure.

Because parcels within the same patient are not independent observations, associations were quantified within each patient and aggregated across the cohort. For each metric, the Spearman correlation with the single-parcel spread reduction was computed across the mPZ parcels of each patient, restricting this within-patient analysis to the 11 patients with at least five mPZ parcels (only patient 2, with four, was excluded), since a rank correlation across fewer parcels is uninformative. The resulting per-patient coefficients were tested against zero with a one-sample Wilcoxon signed-rank test. The pooled estimate was additionally assessed with a permutation test that shuffled the spread reduction within each patient, preserving the patient structure. Partial correlations were used to test whether the associations with the centrality metrics persisted after adjusting for these two dynamical confounds. Finally, each metric was compared between high-impact and low-impact mPZ parcels (single-parcel spread reduction above versus below 0.5).

### 2.8 Minimum replacement set analysis

To dissect the mechanistic origin of WBM-level performance differences between two montages that induce similar field distributions, we developed a per-patient analysis that identifies the smallest subset of cortical parcels driving these differences. Given two montages producing fields *E_n,A_* and *E_n,B_*, with montage A yielding the better seizure-spread outcome in the WBM, the analysis finds the minimum set of parcels at which replacing montage B’s field with montage A’s recovers the better outcome. The categorical composition of this set is a readout of which regions, and which type of correction (additional inhibition vs. reduction of unintended excitation), account for the network-level discrepancy.

The analysis was restricted to parcels where the signed field in A is lower (more inhibitory) than in B (*E_n,A_ < E_n,B_*), since only these can reduce spread when swapped. Within this candidate pool we distinguished three categories: mPZ parcels insufficiently inhibited by montage B (*E_n,B_ <* 0, inhibition must be increased), mPZ parcels actively excited by montage B (*E_n,B_ >* 0, excitation must be decreased), and mNI parcels actively excited by montage B (*E_n,B_ >* 0, excitation must be decreased). EZ parcels were excluded, as our pipeline only considers stimulation effects on the model propagation network.

For each patient, the minimum set was identified in two stages. In the first, we replaced the field at all mPZ candidates and tested whether this alone suppressed spread in the WBM. When it did, an exhaustive combinatorial search identified the smallest mPZ subset achieving suppression. When it did not, mNI involvement is necessary by monotonicity (no subset of mPZ candidates can achieve what the full mPZ replacement cannot), and the analysis proceeded to the second stage.

In the second stage, categorical sufficiency tests were run by replacing the field at all candidates in each category combination (mNI alone, mNI with mPZ-inhibition **ψ**, mNI with mPZ-excitation ↓, and all three) and simulating the WBM, identifying which categories are required to explain the discrepancy in that patient. A heuristic search (iterative parcel removal) then identified a minimum sufficient set within the required categories. This search does not guarantee global minimality, but the categorical composition of its result is constrained by the upstream sufficiency tests and is therefore robust to the search’s limitations.

Finally, parcels for which *E_n,B_ >* 0 and *E_n,A_ <* 0 (ambiguous between an excitation decrease and the need for active inhibition) were disambiguated by simulating the WBM with the parcel’s field set to zero and all other minimum-set parcels unchanged: if spread suppression was retained, the parcel’s contribution was attributed to excitation removal, and otherwise to active inhibition.

## 3 Results

### 3.1 Single-parcel inhibition analysis

First, we investigated the model responses to targeted inhibitory stimulation on single parcels (Section 2.7). As illustrated in Figure 2a, inhibition of an individual parcel could not only prevent seizure activity at the targeted node but also suppress spread throughout interconnected regions. As expected, inhibition of the EZ parcels could reduce seizure spread substantially, but this could also be achieved by inhibiting certain mPZ parcels (non-EZ parcels that seize at baseline in the model).

**Figure 2:**
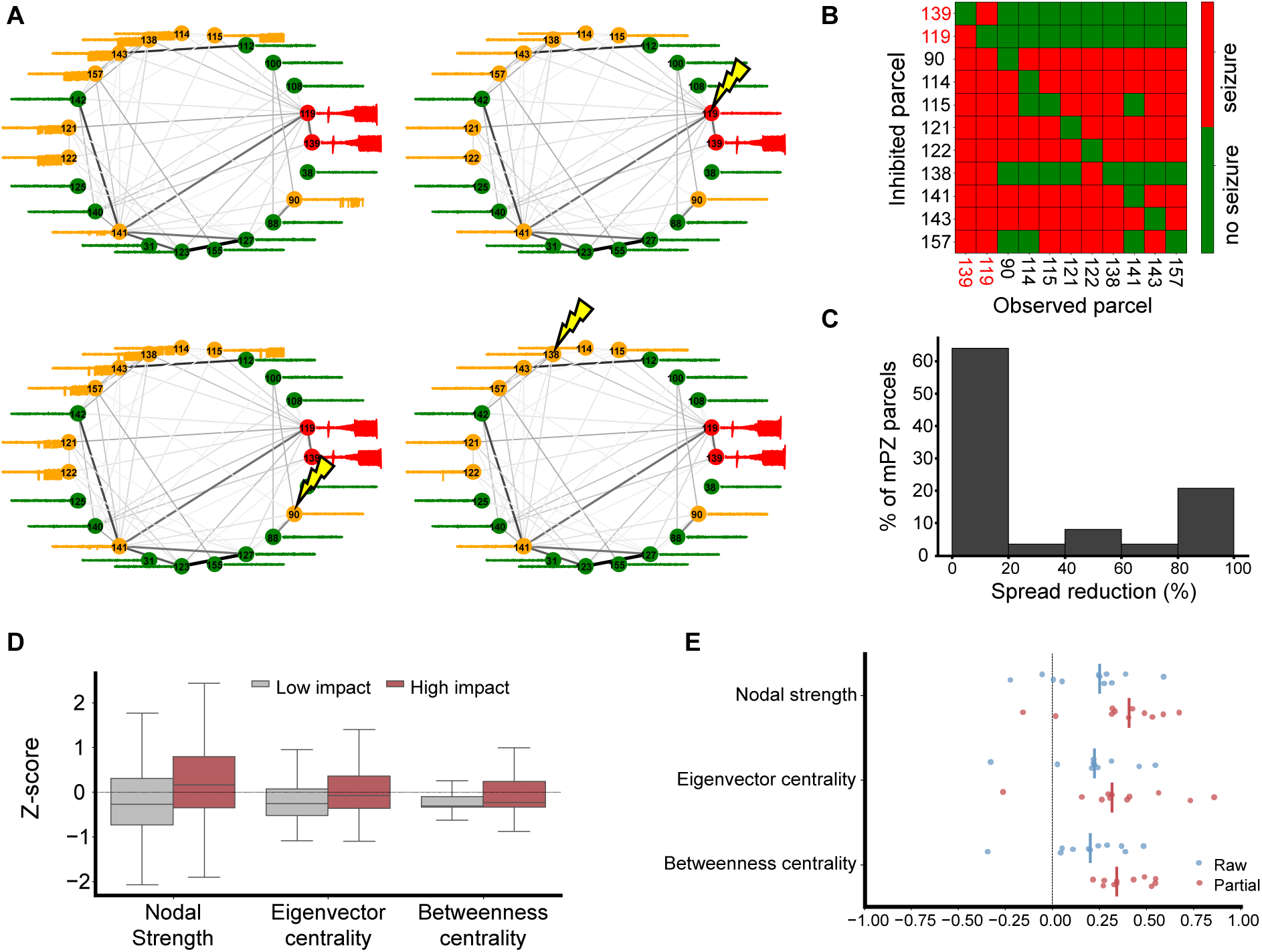
Single-parcel inhibition. a) Synthetic SEEG signals in one of the subject-specific models without stimulation or during inhibitory stimulation at a single parcel (indicated with lightling symbols). Node numbers represent parcel numbers in the VEP parcellation, and edge width represents the connectivity strength between two nodes. Colors indicate the model classification of the regions: red = EZ, yellow = mPZ, green = mNI. SEEG signals are shown without axis labels for clarity, the range displayed is from −1000 to 1000 µV. b) Spread matrix summarizing stimulation effects across all parcels. The y-axis indicates the parcel targeted by inhibitory stimulation, and the x-axis shows the parcel in which seizure activity is observed. Each cell is color-coded to indicate whether seizure activity was present (red) or absent (green). c) Histogram showing the spread reduction that is accomplished by stimulating a single parcel for all subjects and mPZ parcels. d) Network centrality metrics (within-patient z-scored) for mPZ parcels with high (single-parcel spread reduction ≥ 0.5) versus low impact. e) Per-patient Spearman correlations between each network metric and the single-parcel spread reduction, shown as raw correlations and as partial correlations controlling for local excitability (*W*_exc_) and baseline proximity to seizure onset. Each point is one patient and the vertical ticks mark the cohort medians.

To analyze this, we examined the spread matrix for each subject (Figure 2b). The matrix reveals heterogeneity across parcels: inhibition of certain nodes produced widespread suppression of seizures, whereas other nodes had minimal network effects.

We repeated the analysis across all subjects and mPZ parcels. The resulting distribution of spread reduction (Figure 2c) was distinctly bimodal: inhibiting a given parcel typically produced either minimal reduction or near-complete suppression of seizure spread, with few intermediate effects. This pattern is a within-patient phenomenon, not an artifact of a few unusually sensitive subjects: high- and low-impact parcels coexist within individual patients (Figure 2b shows a representative spread matrix). Nor is it an artifact of pooling patients with widely different baseline spread: stratifying the mPZ parcels into patients with generalized (near-whole-brain) versus non-generalized baseline spread, both high- and low-impact parcels were present in each stratum, sharply separated in the generalized cases and more diffusely in the non-generalized ones (Supplementary Figure S2).

Having established that impact is concentrated in specific parcels, we asked whether these high-impact parcels occupy structurally privileged positions in the connectome, relating the single-parcel spread reduction to graph-theoretical centrality metrics computed from each patient’s structural connectome. High-impact parcels were more central than low-impact ones, showing higher nodal strength, eigenvector centrality, and betweenness centrality (Figure 2d). Across mPZ parcels, single-parcel spread reduction was positively associated with all three metrics (per-patient median Spearman *ρ* = 0.20–0.25; Figure 2e and Supplementary Table S1). The associations were consistent in sign across patients (9–10 of 11 for every metric), significant at the patient level (one-sample Wilcoxon *p <* 0.05 for all three), and survived a within-patient permutation test (*p <* 0.001).

These raw correlations could be confounded by the parcels’ dynamical state. Because the model’s dynamical parameters are fitted on top of the structural connectome, the structural metrics may be correlated with how excitable a parcel is or how close it sits to its seizure threshold. Therefore, we recomputed the associations as partial correlations, controlling for each parcel’s excitability (*W*_exc_) and its baseline proximity to seizure onset (defined by how strongly they need to be inhibited to stop seizing). Rather than weakening, the associations strengthened (per-patient median partial |*ρ*| between 0.32 and 0.41; one-sample Wilcoxon *p* ≤ 0.005 for all metrics; Figure 2e). The structural association is therefore genuine rather than an artifact of the parcels’ own dynamical state, which if anything masked it. At the same time, structural position accounted for only a fraction of the variance in single-parcel impact, indicating that the connectome consistently and robustly shapes which parcels are influential without fully determining them. The remaining variance must reflect factors that a static connectome metric cannot capture, such as the dynamical state of the parcels it influences.

### 3.2 Comparison with physics-based pipelines

To compare the field distributions of the montages optimized by the neurotwin and biophysical pipelines, we first computed the *NERNI* of the montages on the target map used in the biophysical optimization (Figure 3a). The field distributions differed substantially. This is expected, since although *NERNI* is part of the loss function, the two pipelines use different target maps.

**Figure 3:**
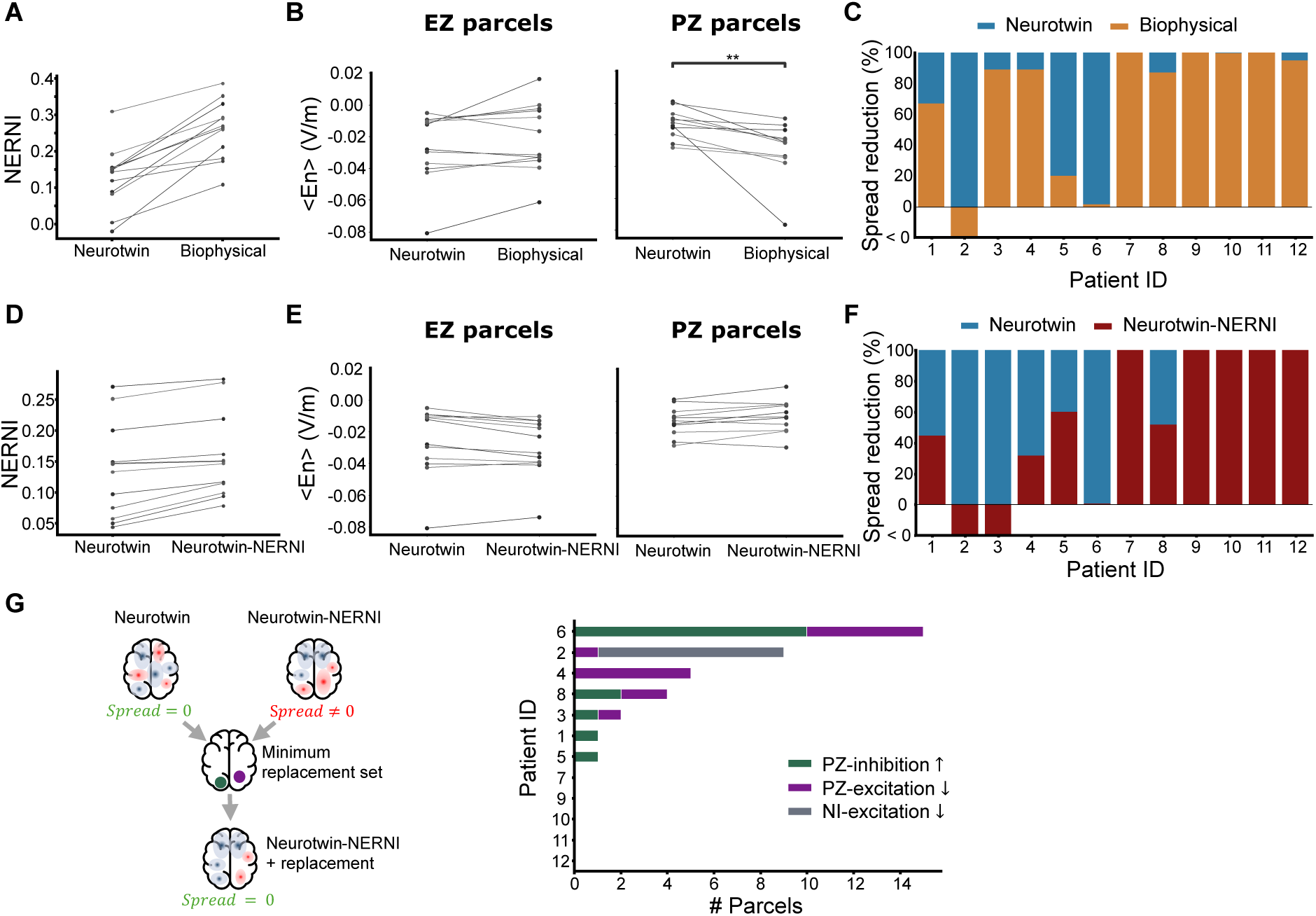
Optimization outputs comparison. a) NERNI of the montages optimized using the neurotwin pipeline and the biophysical model pipeline evaluated in the biophysical pipeline target map. b) Per-subject mean *E_n_*at the EZ and PZ parcels for each pipeline. Each line connects the same patient across pipelines (groups compared with the paired Wilcoxon signed-rank test. ns: no significant difference, *: *p <* 0.05, **: *p <* 0.01). c) Seizure spread reduction in the WBM when the optimized montages are simulated. d-f) Equivalent figures comparing the neurotwin-optimized montages with the montages optimized by minimizing *NERNI* using the target map defined from the WBM. g) Minimum-replacement set analysis. *Left:* schematic illustrating the analysis: for each patient, the minimum set of parcels whose field values must be shifted from the neurotwin-NERNI to the neurotwin solution to recover full spread suppression in the WBM is identified. *Middle:* per-patient composition of the identified minimum sets, color-coded by category: mPZ-inhibition ↑ (mPZ parcels requiring increased inhibition), mPZ-excitation ↓ (mPZ parcels requiring reduced excitation), and mNI-excitation ↓ (mNI parcels requiring reduced excitation).

Figure 3b compares the per-subject mean normal field at clinically defined EZ and PZ parcels between the two pipelines, with each line connecting one patient. At the EZ, the fields did not differ significantly (mean per-subject difference −0.006 V/m, *n* = 12; paired Wilcoxon signed-rank test). At the clinical PZ nodes, the biophysical montages delivered significantly stronger inhibition (per-subject means −0.013 V/m for the neurotwin pipeline and −0.029 V/m for the biophysical pipeline; mean per-subject difference 0.016 V/m, *n* = 11; *p <* 0.01, paired Wilcoxon signed-rank test). This is a moderate difference, attributable to the different target maps (the biophysical pipeline targets EZ and PZ regions equally, whereas the neurotwin pipeline targets the EZ and, with lower weight, the parcels that seize in the WBM) as well as to the network model itself. To place these magnitudes in context, the quantity that drives the WBM is the parcel-averaged normal field *E_n,p_*, which is small in absolute terms because averaging the signed normal component over the folded cortical surface within a parcel produces substantial cancellation. Across all patients, parcels, and optimizations, its absolute value had a median of 0.0132 V/m, and a maximum of 0.1335 V/m.

We then simulated the optimized montages to compare their seizure-spread reduction in the WBM. By construction, the neurotwin pipeline reduces simulated seizure spread to zero in all cases (Figure 3c). The biophysical pipeline, which does not optimize for spread suppression in the WBM, achieved complete spread suppression in 4 cases, near-complete suppression that differed only mildly from the neurotwin montages in 4 further cases, a substantially lower reduction in 3 cases, and in 1 case an increase in seizure spread during stimulation. These results show that despite the significantly stronger PZ inhibition delivered by the biophysical montages, their simulated network-level outcomes differ markedly from those of the neurotwin montages, indicating that, within the model, the overall field magnitude at clinically defined regions is an incomplete predictor of the simulated network response.

Target maps differed between pipelines since one was manually created based on the clinical assessment and the other was defined from the WBM. Thus, the question remains whether the differences between pipelines are solely due to the target map, or whether incorporating the WBM in the optimization has an impact beyond that. To address this, we produced optimized montages using only the *NERNI* term in the loss function (Eq. 6): the target and weight maps were defined from the WBM as explained in Section 2.6.3, and optimization maximized *NERNI*.

As shown in Figure 3d, *NERNI* values were similar between the neurotwin and the “neurotwin-NERNI” optimizations, and the per-subject mean fields differed only marginally (Figure 3e). The mean paired difference was 0.003 V/m at the EZ and −0.003 V/m at the PZ. These magnitudes are negligible: of the order of a few thousandths of V/m and an order of magnitude below the median parcel-average normal field (0.013 V/m).

However, despite the similar field distributions, in the WBM (Figure 3f) the neurotwin-NERNI montages produced markedly lower simulated spread reductions in 5 subjects and increased spread in 2 cases. Because both arms share the same WBM-derived target map and differ only in whether the network term enters the loss, this divergence isolates the WBM as the factor that changes the resulting montages and their simulated outcomes.

The previous results showed that neurotwin and neurotwin-NERNI montages can produce very similar field distributions yet yield markedly different seizure-spread outcomes in the WBM. To investigate the mechanistic origin of this discrepancy, we identified, for each patient, the minimum set of parcels whose field values must shift from the neurotwin-NERNI to the neurotwin solution to recover full spread suppression (see Section 2.8 and Figure 3g). The categorical composition of these sets provides a readout of how the two optimizations diverge mechanistically: which parcels were under-inhibited, which were over-excited, and whether the relevant parcels lay within or outside the model propagation network (mPZ vs mNI).

Of the 12 patients, 7 showed a spread discrepancy between the two pipelines and were included in this analysis. The remaining 5 achieved full suppression with both pipelines and were not analyzed (Figure 3g, middle). Across the included patients, minimum sets were often small: 5 of 7 patients required ≤ 5 parcels, with patients #2 and #6 the exceptions, requiring 9 and 15 respectively (where the heuristic second-stage search was used, these set sizes are upper bounds on the true minimum; see Section 2.8). Although the two solutions do not differ significantly in their average EZ or PZ fields (above), the field changes at the specific minimum-set parcels were larger: the absolute field difference at these parcels (median 0.013 V/m) was of the same order as the parcel-average fields themselves (cf. Figure 3b). The divergence in network outcome therefore does not stem from a global difference in the delivered field but from changes concentrated at a limited number of specific parcels.

The categorical composition of these minimum sets reveals two complementary mechanisms by which the neurotwin-NERNI montages did not achieve full spread suppression. Pure mPZ involvement explained the discrepancy in 6 of 7 patients, with the remaining patient (#2) requiring substantial reduction in mNI excitation (8 of 9 parcels in the minimum set). Within the patients explained by mPZ alone, the under-inhibition mechanism (mPZ-inhibition ↑) and the over-excitation mechanism (mPZ-excitation ↓) appeared with comparable frequency: under-inhibition in 5 of 7 patients, over-excitation in 5 of 7, with 3 patients requiring both. In other words, the different performance of the montages can be explained approximately as often by leaving seizure-relevant mPZ regions insufficiently inhibited as by actively exciting mPZ regions that should have been spared.

These findings highlight a conceptual limitation of conventional biophysics-based optimization: such pipelines tend to treat all targeted regions (clinical EZ and PZ), and all non-target regions, as homogeneous within their respective groups. The WBM-based analysis, however, reveals heterogeneity along two dimensions. First, among the parcels that actually propagate seizures in the model (mPZ), not all contribute equally to network spread: some act as critical hubs whose inhibition determines whether spread is suppressed (consistent with the single-parcel inhibition results of Section 3.1), while over-excitation of others can actively worsen network outcomes. Second, parcels outside the model propagation network (mNI) are not uniformly inconsequential: occasionally sensitive parcels can worsen spread when excited. Together, these results suggest that effective non-invasive seizure control may require optimization strategies that explicitly account for heterogeneous network roles of the different regions.

Since the WBM’s sensitivity to stimulation is governed by the electrical coupling gain *G_E_*, we verified that the above findings do not depend on its calibration by repeating the optimization with *G_E_*doubled and halved (reference fields of 0.05 and 0.2 V/m; Supplementary Figure S3).

The optimal montage was gain-dependent: neurotwin montages optimized at the primary calibration no longer fully suppressed spread when re-evaluated at the doubled or halved gain in 7 and 5 of 12 patients, respectively. However, after re-optimizing we recovered complete suppression in all patients at both calibrations and the re-optimized montages remained close to the primary-calibration ones in terms of their field distributions as shown by the NERNI changing by a mean of only 0.006 (maximum 0.034) across patients and calibrations. The minimum-replacement-set analysis between the primary-calibration and re-optimized montages localized this difference to a small number of parcels, of the same mechanistic categories identified above. Halving the gain required correcting at most two parcels per patient, whereas doubling it required somewhat more (four and seven parcels in two patients) and consisted predominantly of reductions of excitation at mPZ parcels. This asymmetry is consistent with the more sensitive, higher-gain model amplifying the montage’s incidental excitatory effects, the same off-target excitation mechanism identified above. In every case, however, the prescription’s gain sensitivity remained confined to a subset of seizure-relevant parcels rather than a global change in the delivered field.

Crucially, the divergence from the field-only optimization persisted throughout: the neurotwin-NERNI montages failed to reach full or near-complete suppression in 6 of 12 patients at the doubled gain and 8 of 12 at the halved gain (7 of 12 at the primary calibration), and actively increased seizure spread in 2 and 1 patients, respectively (2 at the primary calibration). The divergence did not collapse toward convergence even when *G_E_* was doubled (the regime in which an over-sensitive model would render the network term uninformative), indicating that the discriminative power of the network objective is not an artifact of the chosen gain.

### 3.3 Retrospective analysis

Six patients from the present dataset participated in an open-label clinical trial in which tDCS was administered in three stimulation cycles (2 × 20 minutes per day for five consecutive days),^15^ with seizure frequency during and after treatment compared to baseline. As throughout this manuscript, these patients are referred to by their present-study IDs (#4–#9). Their correspondence with the IDs used by Bartolomei et al.^15^ (prefixed B) is: #4↔B1, #5↔B2, #6↔B4, #7↔B7, #8↔B9, and #9↔B14. Two patients met the predefined responder criterion (*>*50% reduction in seizure frequency), while four were classified as non-responders. Per-patient seizure frequency changes relative to baseline are shown in Figure 4a (left).

**Figure 4:**
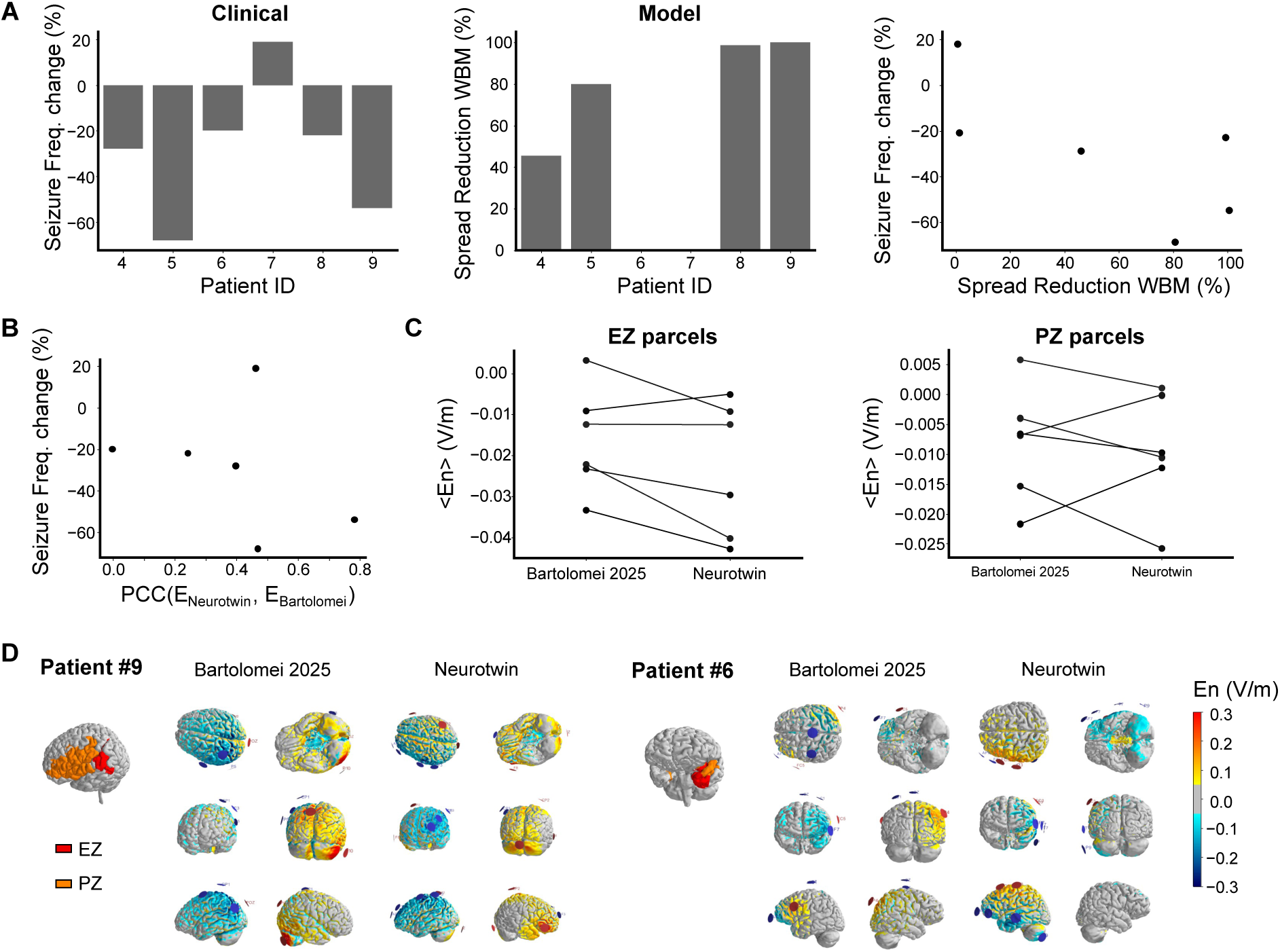
Retrospective validation against clinical outcomes. a) Comparison of clinical and model-predicted outcomes. Left: Seizure frequency change relative to baseline in the six patients during the clinical trial of Bartolomei et al. (2025).^15^ Patient IDs follow the numbering used in the present study (#4–#9). Center: percentage reduction in seizure spread predicted by the WBM when the clinically applied montages are simulated in each patient’s neurotwin model. Right: relationship between clinical seizure frequency change and model-predicted spread reduction across patients (Spearman *ρ* = −0.71, *p* = 0.11). b) Clinical seizure frequency change as a function of the Pearson’s correlation coefficient (PCC) between the field distributions induced by the neurotwin-optimized montages and clinically applied montages (Spearman *ρ* = −0.60, *p* = 0.21). c) Average normal component of electric field induced by the neurotwin-optimized montages and the montages applied in the clinical trial at the clinically defined epileptogenic zone (EZ) and propagation zone (PZ) parcels. Each point is one patient’s mean field and lines connect the same patient across montages. No statistically significant differences were found between groups (paired Wilcoxon signed-rank test). d) Electric field distributions (*E_n_*) induced by each montage in the cases with highest (#9, PCC = 0.78) and lowest (#6, PCC = 0.00) correlations between the field distributions. For each patient, the EZ and PZ areas are displayed on the gray matter surface.

To assess whether the neurotwin framework would have predicted these clinical outcomes, we simulated each patient’s clinically applied montage (the exact trial montage, designed with the same biophysical head-model approach used here) in their corresponding neurotwin and quantified the resulting reduction in seizure spread in the WBM. Across the six patients, predicted spread reductions ranged from 0% to nearly 100% (Figure 4a, center), broadly tracking the heterogeneity observed in the clinical data. When the two readouts were compared directly (Figure 4a, right), patients with greater WBM-predicted spread reduction tended to show larger reductions in clinical seizure frequency (Spearman *ρ* = −0.71, *p* = 0.11, *n* = 6). Because seizure-frequency change is signed, with a reduction counting as negative, while spread reduction is a positive percentage, a beneficial relationship appears as a negative correlation. This correlation did not reach statistical significance, which is unsurprising given the small sample, and should not be interpreted as evidence of predictive accuracy. Nonetheless, both the magnitude of the rank correlation and the visual trend are directionally consistent with the hypothesis that WBM-predicted spread reduction relates to clinical response.

We next asked whether the spatial similarity between each patient’s clinically applied montage and their neurotwin-optimized montage was associated with clinical outcome. For each patient, we computed the Pearson correlation coefficient (PCC) between the field distributions induced by the two montages. Patients whose clinical montage produced a field distribution more similar to the neurotwin-optimized solution tended to experience larger reductions in seizure frequency (Spearman *ρ* = −0.60, *p* = 0.21, *n* = 6; Figure 4b). The relationship is in the expected direction but weaker than the association with the WBM-predicted spread reduction reported above. These two readouts are related but not equivalent, and the difference between them is itself informative. The field-distribution PCC is a global measure of spatial similarity that weights all parcels equally. As the minimum-replacement-set analysis shows, however, two montages with highly correlated field distributions can still differ at a small number of seizure-relevant parcels and thereby produce very different seizure spread in the WBM. The WBM-predicted spread reduction integrates precisely these localized, network-weighted differences to which the global PCC is largely insensitive. In principle, this makes the WBM-predicted spread reduction a more sensitive readout of clinical response than the global field similarity, in line with the central finding of this study that the network response, rather than the aggregate field, is what distinguishes montages. With *n* = 6, however, the two correlations are statistically indistinguishable and neither is significant, so we do not interpret the difference between them.

One patient in this retrospective subset (#9) is a high-leverage point for these correlations: it was both a clinical responder and the patient whose clinically applied montage produced near-complete spread reduction in the WBM, yet it has the poorest functional-connectivity fit of the subset (Table 2). Because such a point could disproportionately drive the association while resting on a model of questionable validity, we repeated both analyses after excluding it to test whether the correlations hinge on this poorly-fit case. The WBM-predicted spread reduction remained essentially unchanged (*ρ* = −0.70, *p* = 0.19, *n* = 5), indicating that this association is not driven solely by the poorly-fit patient, while the field-similarity association was more affected (*ρ* = −0.40, *p* = 0.50, *n* = 5). At *n* = 5, however, a single rank change moves either coefficient substantially, so we do not read this difference as evidence that one readout is more robust than the other.

Consistent with the results in the previous section, where biophysics-based and neurotwin-optimized montages delivered comparable EZ fields (differing only moderately at the PZ) despite differing network-level effects, no statistically significant differences were found between the clinical and neurotwin-optimized montages in the per-subject mean normal field at EZ or PZ parcels (Figure 4c; paired Wilcoxon signed-rank test). The two patients with the highest and lowest PCC values (#9, PCC = 0.78; #6, PCC = 0.00) are shown in Figure 4d, illustrating the spatial range of agreement between the clinical and neurotwin-optimized montages: in patient #9 the field distributions are broadly similar across the cortex, whereas in patient #6 they differ substantially in both magnitude and location.

These correlations are encouraging but preliminary. They raise the possibility that the neurotwin framework could help explain the heterogeneous response to tDCS observed in previous clinical trials. However, with only six patients none of the reported correlations reaches significance and no multiple-comparison correction was applied, so these results are strictly hypothesis-generating and cannot support any firm claim.

## 4 Discussion

In this study, we introduced a whole-brain modeling-based optimization pipeline for tDCS in drug-resistant epilepsy and compared the montages it produces with those of conventional physics-based approaches. Because the neurotwin pipeline optimizes simulated seizure spread directly, it reduces spread to zero in the WBM by construction. The informative finding is not this suppression but the observation that, even though the local electric field distributions at the clinically defined EZ are comparable across pipelines (with only a moderate difference at the PZ), the network-informed and field-based optimizations yield substantially different montages with markedly different simulated network outcomes. This dissociation is necessarily a within-model result: because the field-based pipelines do not optimize simulated spread, the divergence does not by itself establish that the neurotwin montages are clinically superior. While conventional pipelines optimize tDCS montages to maximize the inhibitory field of predefined cortical targets, the neurotwin pipeline incorporates the epileptogenic network dynamics. We hypothesize that accounting for these dynamics could translate into more effective seizure control, a hypothesis that remains to be tested prospectively.

We next compared the full neurotwin optimization with the neurotwin-NERNI variant, which shared the same WBM-derived target map and differed only in whether the network dynamics entered the loss function. The two produced essentially matched field distributions yet diverged in their simulated network outcomes, isolating the whole-brain model itself, rather than the choice of target map, as the source of the difference. The minimum-replacement-set analysis localized this divergence to a small number of seizure-relevant parcels and revealed two complementary failure modes of field-only optimization: insufficient inhibition of critical mPZ parcels and active over-excitation of parcels that should have been spared, in comparable measure. The aggregate field delivered to clinically defined targets is therefore an incomplete predictor of the network response, and where in the network the field is delivered is equally decisive. These findings suggest that a guiding assumption of conventional optimization, namely that inhibiting the target areas while sparing the rest maximizes efficacy and that all targeted regions can be treated alike, may be incomplete: the parcels whose inhibition most reduces spread instead occupy integrative, highly connected positions in the structural connectome, an association that persisted, indeed strengthened, after accounting for their local excitability and proximity to seizure onset.

The fact that the connectome position predicts single-parcel impact consistently across patients shows the effect is partly grounded in anatomy rather than an artifact of the fitted models. However, the correlation is moderate, showing that the connectome is necessary but not sufficient and reinforcing the need for the personalized WBM. The unexplained variance is not due to the parcels’ own dynamical state, for which we controlled, but more likely to how strongly a parcel drives the other seizing parcels and to how close those parcels sit to their own threshold, a structural-dynamical combination that whole-connectome metrics cannot capture. Consistent with this, single-parcel impact is a one-at-a-time measure, whereas suppression often depends on sets of parcels acting together, as the minimum-replacement-set analysis shows.

Our results also suggest that some parcels are sensitive to stimulation in the opposite direction: exciting them can worsen seizure spread. Minimizing stimulation outside the target may therefore not be enough, and some non-target regions must be carefully managed to maximize treatment benefit.

From a clinical perspective, this has direct relevance for understanding heterogeneous patient responses to tDCS. Patients whose seizure dynamics critically depend on network hubs outside the EZ may fail to respond, or even worsen, when montages are optimized solely on the basis of EZ targeting. This network-level mechanism could help explain why some individuals in prior tDCS trials showed limited benefit despite apparently successful inhibition of EZ regions.^12, 13, 15^ By explicitly evaluating how fields in both EZ and non-EZ regions influence seizure propagation, the neurotwin framework may provide a more complete and mechanistically grounded rationale for montage design.^71^

Our proposed framework is closely related to the digital-twin lineage of The Virtual Brain and the Virtual Epileptic Patient,^24, 31, 72^ but is tailored for tES. Whereas such models have mostly guided diagnosis or surgical planning,^27, 32, 35, 36, 38^ and recent stimulation work has served diagnostic ends,^45^ here the simulated network response is the objective that drives non-invasive montage design. In this way, the present work extends the digital-twin paradigm in epilepsy from diagnostic and surgical applications towards the design of non-invasive neuromodulation therapy. Conversely, the mechanistic insights may feed back into the surgical applications from which this paradigm originated: that decisive nodes can lie in interconnected hubs beyond the clinically defined EZ offers a candidate explanation for why EZ-focused resections sometimes fail, and, together with the finding that perturbing individual nodes does not uniformly reduce spread, could help refine network-guided minimally invasive procedures such as SEEG-guided thermocoagulation.^73^

Our modeling framework relies on the imprint hypothesis, assuming that acute effects of stimulation are indicative of long-term plastic outcomes. While supported by some experimental evidence,^5, 59^ and consistent with evidence from invasive neuromodulation that stimulation-induced network changes track long-term seizure outcomes,^74^ this remains to be further validated, especially for the specific tDCS application in epilepsy. Integrating plasticity rules in the modeling framework would enable simulations of long-term outcomes and optimizing the stimulation schedule (e.g., number of sessions and duration). However, it is unclear whether this would impact the optimal electrode montage. Relatedly, our pipeline takes the reduction of acute seizure propagation as its optimization objective. This rests on the imprint hypothesis together with the premise that limiting propagation is therapeutically beneficial. Alternative objectives, such as reducing the intrinsic excitability of the epileptogenic network or targeting cumulative plastic change, are also plausible and they could even be combined with spread suppression in a single multi-objective loss. Future work should examine such approaches.

Moreover, the WBM framework presented here depends on parameters that are not tightly constrained, most notably the coupling gain *G_E_*. A sensitivity analysis over a fourfold range of *G_E_* (Supplementary Figure S3) showed that the divergence from field-only optimization persists and that *G_E_* uncertainty produces only a bounded, localized change in the prescribed montage, so the central findings do not hinge on the calibration. A related limitation concerns the propagation network itself. Both the clinically defined PZ and the model-defined mPZ are phenomenological constructs, and there is currently no validated, quantitative signal-based criterion for the true extent of a patient’s propagation zone against which to benchmark them. This makes the boundary of the propagation network partly model-dependent, as reflected in its wide range across patients.

In the present study, we compared the neurotwin approach with conventional pipelines through *in silico* analyses and a small retrospective analysis. Therefore, further validation is essential. The pipeline presented here is currently being deployed in a clinical trial (NCT06334952) to assess its safety and efficacy. However, besides this prospective validation, future work should also focus on a more thorough retrospective validation of neurotwin-optimized montages by extending the retrospective analysis to larger clinical datasets including patients with known clinical outcomes after tDCS.

Finally, an important limitation of the framework as currently implemented is its dependence on SEEG: both the EZ/PZ classification and the functional-connectivity fitting of the WBM require intracranial recordings. This restricts the approach, in its present form, to patients undergoing pre-surgical SEEG evaluation. Future work should focus on developing neurotwin personalization pipelines driven by non-invasive recordings such as scalp EEG or MEG: the present SEEG-based pipeline is a step towards that objective, establishing the optimization framework and the mechanistic principles on which a fully non-invasive version would build, and which would in turn extend the approach to a substantially broader patient population.

## 5 Conclusions

We introduced a neurotwin pipeline for the optimization of personalized multichannel tDCS in drug-resistant epilepsy, integrating a biophysical head model with a personalized whole-brain model of seizure dynamics. Although the montages it produced delivered comparable inhibitory fields at the clinically defined EZ, and a moderately weaker inhibitory field at the PZ, relative to conventional biophysics-based optimization, the two approaches yielded substantially different montages with different simulated network outcomes. Because the whole-brain modeling pipeline optimizes simulated spread directly, it is this divergence, rather than the suppression itself, that constitutes the informative result. The divergence persisted when the target map was held fixed, showing that embedding network dynamics directly in the optimization loss, and not only in the target-map definition, changes the resulting montages. Mechanistically, the difference could be traced to the heterogeneous roles that individual cortical regions play in seizure propagation, namely critical propagation-zone parcels whose inhibition is decisive and sensitive off-target parcels whose excitation can be detrimental. These are features that field-only optimization cannot capture. These decisive propagation-zone parcels tended to be the integrative, highly connected hubs of each patient’s structural connectome. A preliminary retrospective analysis showed that model-predicted spread reduction tracked clinical seizure-frequency change in the expected direction, though the correlation was not statistically significant in this small sample.

## Data Availability

The data used for this manuscript are not publicly available because the patients did not consent for the sharing of their clinically obtained data. Requests to access to the datasets should be directed to the corresponding author.

## 6 Acknowledgements

This work has received funding from the European Research Council (ERC) under the European Union’s Horizon 2020 research and innovation programme (Grant Agreement No. 855109; ERC-SyG 2019) and from FET under the European Union’s Horizon 2020 research and innovation programme (Grant Agreement No. 101017716).

## Contributions

**B. Mercadal**: Conceptualization, Methodology, Software, Formal analysis, Investigation, Writing – original draft. **E. Lopez-Sola**: Methodology, Software, Formal analysis, Writing – review & editing. **J. Makhalova**: Resources, Data curation, Writing – review & editing. **F. Pizzo**: Resources, Data curation, Writing – review & editing. **R. Salvador**: Methodology, Software, Writing – review & editing. **F. Bartolomei**: Resources, Data curation, Funding acquisition, Writing – review & editing. **G. Ruffini**: Conceptualization, Supervision, Funding acquisition, Writing – review & editing.

