## Supplementary Materials for "A whole-brain modeling framework for tDCS montage optimization in drug-resistant epilepsy"

### Supplementary Methods: whole-brain model personalization

This section summarizes the whole-brain model (WBM) personalization procedure of Lopez-Sola et al.[1] on which the present pipeline builds, so that the modeling layer can be followed without consulting the original paper. All choices below follow[1] unless stated otherwise.

**Network and node models.** A neural mass model (NMM) is placed at each of the 162 cortical parcels of the VEP atlas,[2] and two NMM types are used. EZ parcels use the seizure-generating NMM of Lopez-Sola et al.,[3] which incorporates chloride dynamics in the SST→P and PV→P synapses and reproduces the transition from interictal to fast-onset to clonic activity. In these nodes the seizure is triggered deterministically by an external input of 110 Hz delivered to the pyramidal population at $t = 15$ s. All other parcels (clinical PZ and NIZ) use a Wendling-class model[4] with parameters taken from,[3] in which crossing a saddle-node bifurcation produces a large-amplitude periodic (ictal) regime. The mean external input to these nodes is fixed at $\mu = 15$ Hz, just below the bifurcation threshold even at the maximum excitability tested, so that a non-EZ node can enter ictal dynamics only when it receives sufficient input from other active nodes. Seizure propagation in the model is therefore driven by network interactions rather than by isolated nodes seizing spontaneously.

**Coupling.** Nodes are coupled through the patient's structural connectome, obtained from diffusion-MRI tractography (20 million streamlines, normalized by the total streamline count divided by the number of parcels). Long-range excitatory input reaches each pyramidal population in proportion to the connectome weight scaled by a global coupling gain $G$ shared across the network. $G$ is restricted to $[1, 300]$ to avoid both disconnection of the network and saturation of the neural masses.

*

**Fitted parameters.** Personalization fits, for each patient: (1) the global coupling gain $G$; (2) the excitatory synaptic gain $W_{\mathrm{exc}}$ of each parcel sampled by SEEG, a proxy for regional excitability, restricted to $[3, 10]$ mV; and (3) a single gain $W^*_{\mathrm{exc}}$ applied to all parcels not sampled by SEEG. The lower bound of 3 mV is treated as a healthy baseline (the nearest integer to the Jansen-Rit value of 3.25 mV), and the upper bound of 10 mV is the point beyond which further increases no longer raise the probability of entering ictal dynamics. $W_{\mathrm{exc}}$ is fitted only for SEEG-sampled parcels because the empirical FC constrains only those parcels. Fitting it for unsampled parcels has negligible effect on the synthetic FC and introduces optimization degeneracy, so a single $W^*_{\mathrm{exc}}$ captures unsampled excitability instead. The fitted and fixed quantities are listed in Table .

**Empirical functional connectivity.** For each parcel sampled by SEEG, one bipolar signal is selected by prioritizing epileptogenicity (EZ before PZ before NIZ) and then the highest seizure-to-baseline energy ratio. For each seizure, a window from 10 s before to 20 s after the onset of the clonic phase is extracted. Each signal is band-pass filtered (0.5 Hz to the Nyquist frequency), its amplitude envelope is computed as the modulus of the Hilbert transform and low-pass filtered below 1 Hz, and the result is normalized to unit standard deviation and demeaned. The functional connectivity between two parcels is the maximum of the absolute cross-correlation of their envelopes, which accommodates propagation delays. For patients with multiple recorded seizures, the empirical FC is the mean over seizures.

**Simulated SEEG and functional connectivity.** Synthetic SEEG is generated with the laminar neural mass modeling framework[5] and the laminar forward model of Mercadal et al.:[6] the average post-synaptic currents into pyramidal cells drive a current-source-density profile within a cortical patch, from which the voltage recorded by a bipolar contact pair is derived. Synthetic FC is then computed from these synthetic signals with the same amplitude-envelope procedure used for the empirical data, ensuring a like-for-like comparison.

**Loss function and optimization.** The parameters are fitted by minimizing

$$L = -\,\mathrm{PCC}(\mathrm{FC}_{\mathrm{sim}}, \mathrm{FC}_{\mathrm{emp}}) + R, \tag{1}$$

where PCC is the Pearson correlation between the simulated and empirical FC matrices and $R$ is an $L_1$ regularization term that penalizes deviations of $W_{\mathrm{exc}}$ from the healthy baseline $W_{\mathrm{exc},h} = 3$ mV,

$$R = \frac{1}{N_{\mathrm{SEEG}} + 1} \sum_i \lambda \left| W_{\mathrm{exc},i} - W_{\mathrm{exc},h} \right|, \tag{2}$$

the sum running over the $N_{\mathrm{SEEG}}$ sampled parcels plus the single non-sampled gain $W^*_{\mathrm{exc}}$, and $\lambda$ setting the regularization strength. This term reflects the assumption that most regions are close to healthy excitability and only a few are pathologically hyperexcitable. The loss is minimized with the differential evolution algorithm[7] (SciPy `best1bin` strategy[8]), searching over integer parameter values, with $G$ initialized uniformly on its range and $W_{\mathrm{exc}}$ initialized from a half-Laplacian distribution centered at the healthy baseline. Each candidate model is simulated for 50 s at 1000 Hz with a fourth-order Runge-Kutta solver.

**Model validity.** A fitted model is retained only if it lies in a physiologically plausible regime, following:[1] (1) a seizure is present in all EZ nodes; (2) the clonic phase in EZ nodes does not begin before 5 s; (3) no PZ or NIZ node seizes before the EZ clonic phase begins; and (4) the difference between the minimum simulated and the minimum empirical FC does not exceed 0.2, which rules out spurious high-correlation fits in which all nodes seize. In addition, as reported in the main text, the fitted FC correlation must exceed the correlation between the empirical FC and the structural connectome, confirming that the model captures dynamics beyond what the anatomy alone encodes.

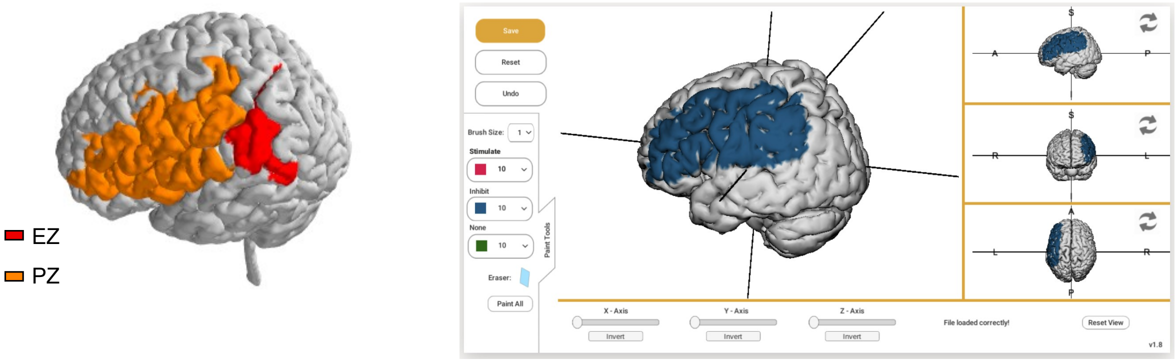


Figure S1: **Definition of the clinician-based target map for the biophysical optimization pipeline.** Illustration of the target-map painting step used in the biophysics-based optimization (Figure 1b) for one representative patient. **(left)** Epileptogenic-zone (EZ) and propagation-zone (PZ) parcels, as classified by the clinician from the SEEG evaluation, displayed on the gray-matter surface. **(right)** The corresponding inhibitory target map painted for the same patient in the online target editor, marking the cortical regions to be inhibited. This clinician-defined target map is the input to the biophysical (field-based) optimization and is distinct from the target map derived from the WBM used in the neurotwin pipeline.

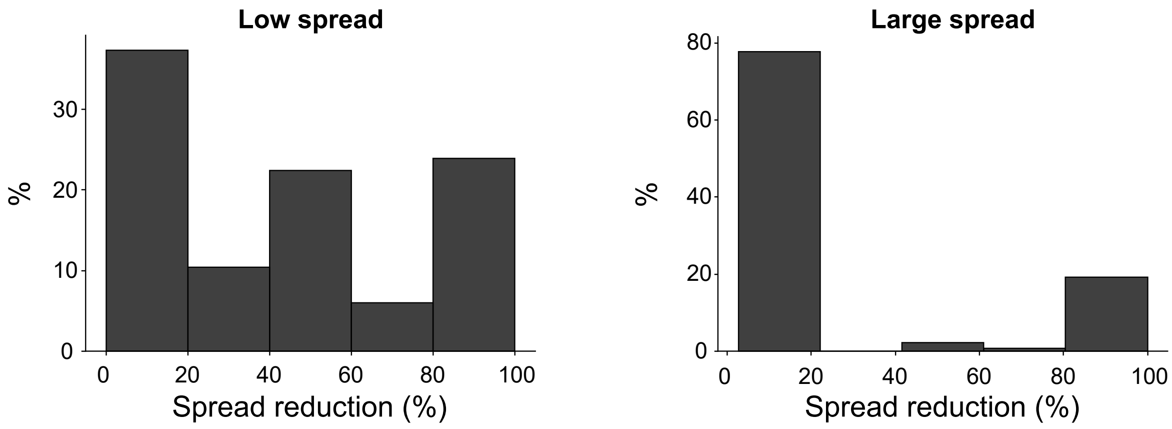


Figure S2: **Single-parcel spread reduction stratified by baseline spread.** The distribution of single-parcel spread reduction (as in Figure 2c) is shown separately for mPZ parcels of patients with low baseline spread (left) and large, near-whole-brain baseline spread (right). In the large-spread stratum the distribution is sharply bimodal, with parcels producing either minimal reduction or near-complete suppression and almost none in between. In the low-spread stratum the same two extremes are present, a dominant low-reduction peak together with a distinct subset of high-impact parcels, though with more intermediate values. High- and low-impact parcels therefore coexist in both strata, confirming that the bimodality reported in Figure 2c is not an artifact of pooling patients across the wide range of baseline spread reported in Table 2.

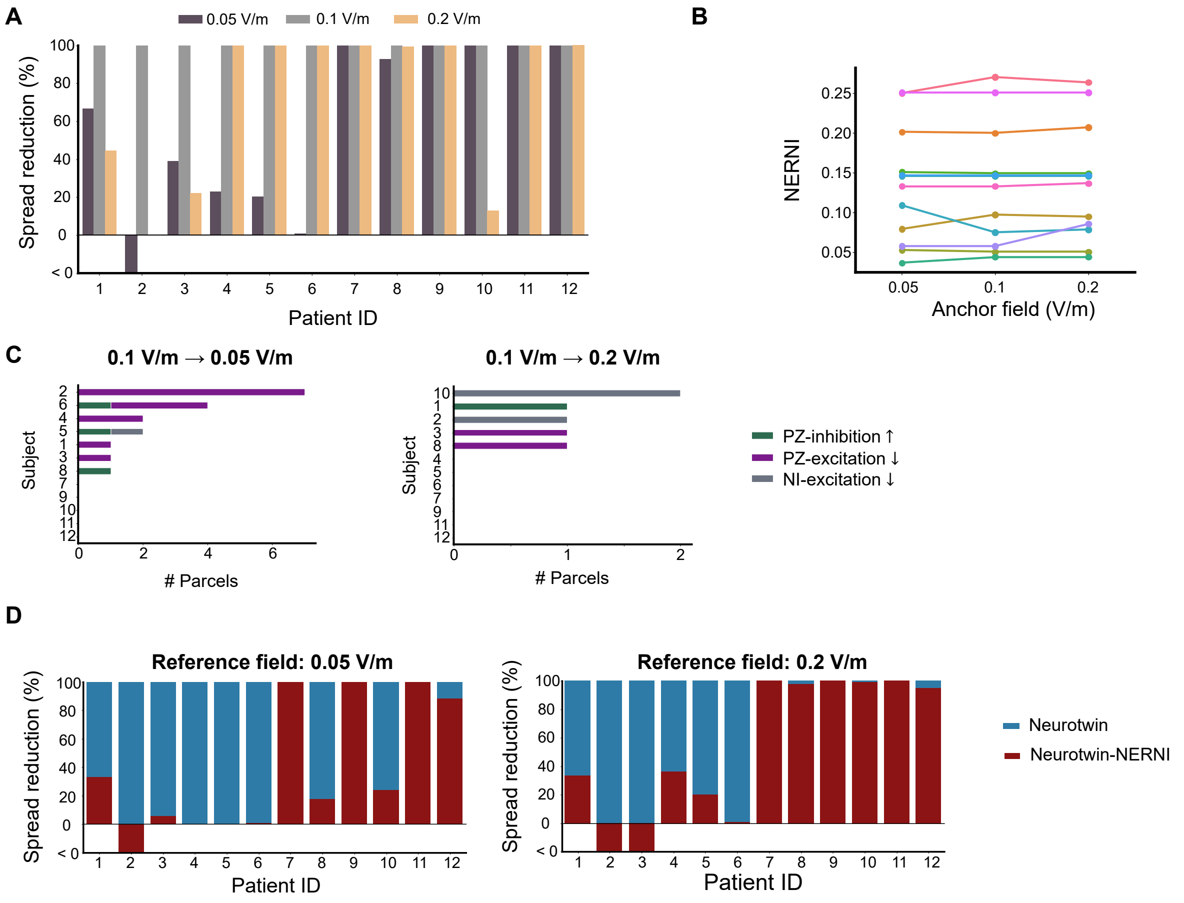


Figure S3: **Robustness to the electrical coupling gain $G_E$.** The optimization was repeated with $G_E$ doubled and halved relative to the primary calibration (reference fields of 0.05, 0.1, and 0.2 V/m). **(a)** Seizure-spread reduction of the neurotwin montages optimized at the primary calibration (0.1 V/m) when applied at each of the three gains. Values below 100% identify the patients in whom the montage no longer fully suppressed spread and was re-optimized (7 patients at 0.05 V/m, 5 at 0.2 V/m). **(b)** NERNI of the re-optimized montages on the WBM-derived target map, per patient and gain, showing that the field-matching quality changes little across calibrations. **(c)** Categorical composition of the minimum-replacement set between the primary-calibration and re-optimized montages at 0.05 and 0.2 V/m: the number of parcels, and their type (increased mPZ inhibition, reduced mPZ excitation, or reduced non-involved excitation), that account for the montage change in each patient. **(d)** Seizure-spread reduction in the WBM for the neurotwin and neurotwin-NERNI montages at 0.05 and 0.2 V/m. Across this fourfold range the neurotwin pipeline recovered full suppression at every calibration, its divergence from the field-only optimization persisted and did not collapse at the higher gain, and the difference between primary-calibration and re-optimized montages remained confined to a small number of seizure-relevant parcels.

Table S1: **Network-centrality analysis of single-parcel impact.** Per-patient Spearman correlations between each centrality metric and the single-parcel spread reduction, computed across mPZ parcels and aggregated over the $n = 11$ patients with at least five mPZ parcels. Median $\rho$ and the one-sample Wilcoxon signed-rank test are taken across patients. "Consistent sign" is the number of patients whose correlation matched the cohort-median sign. Partial correlations control for local excitability ($W_{\text{exc}}$) and baseline proximity to seizure onset. For every metric the pooled association was additionally confirmed by a within-patient permutation test ($p < 0.001$).

| Network metric | Median $\rho$ | Consistent sign | Wilcoxon $p$ | Partial median $\rho$ | Partial Wilcoxon $p$ |
|---|---|---|---|---|---|
| Nodal strength | 0.25 | 9/11 | 0.019 | 0.41 | 0.003 |
| Eigenvector centrality | 0.22 | 10/11 | 0.032 | 0.32 | 0.005 |
| Betweenness centrality | 0.20 | 10/11 | 0.024 | 0.34 | $< 0.001$ |

Table S2: **Fitted and fixed parameters of the whole-brain model personalization**.[1] Fitted parameters are optimized per patient to match empirical functional connectivity; fixed parameters are held constant across patients. Node-level NMM parameters not listed here (for example the chloride-dynamics parameters of the EZ model) are taken unchanged from.[3]

| Parameter | Symbol | Role | Value / range |
|---|---|---|---|
| Global coupling gain | $G$ | fitted (per patient) | $[1, 300]$ |
| Excitatory synaptic gain, SEEG-sampled | $W_{\text{exc}}$ | fitted (per parcel) | $[3, 10]$ mV |
| Excitatory synaptic gain, non-sampled | $W^*_{\text{exc}}$ | fitted (single value) | $[3, 10]$ mV |
| Healthy excitability baseline | $W_{\text{exc},h}$ | fixed | 3 mV |
| External input to PZ/NIZ pyramidal cells | $\mu$ | fixed | 15 Hz |
| EZ seizure-trigger input | – | fixed | 110 Hz at $t = 15$ s |
| Simulation duration / sampling rate | – | fixed | 50 s / 1000 Hz (RK4) |
